# Age-related structural remodelling of the coronary microcirculation

**DOI:** 10.1101/2024.02.27.24303474

**Authors:** Daniel Faria, Marco Lombardi, Nina van der Hoeven, Alejandro Travieso, Julius C. Heemelaar, Sukhjinder S. Nijjer, Hernán Mejía-Rentería, Guus A. de Waard, Sayan Sen, Tim P van de Hoef, Ricardo Petraco, Mauro Echavarría-Pinto, Jan J Piek, Justin E Davies, Niels van Royen, Javier Escaned

## Abstract

**Background:** While it is broadly accepted that ageing is associated with impairment of coronary microvascular function, little is known on the underlying mechanisms. Diastolic microcirculatory conductance (DMVC) and the backward expansion wave (BEW) derived from wave intensity analysis (WIA) are two physiological indices derived from post-analysis of coronary pressure and flow that have been previously validated against endomyocardial biopsy micromorphometry, thus serving as metrics of structural microcirculatory remodelling applicable to in vivo assessment of the human coronary microcirculation. We investigated age-related changes in coronary microvascular structure in patients with stable angina without epicardial coronary stenoses.

**Methods:** This is an analysis of the IDEAL registry, including a total of 165 vessels without coronary stenosis interrogated with combined pressure/Doppler guidewires in non-diabetic patients. We calculated DMVCs and BEWs using dedicated software, and we compared them between patient groups according to age tertiles. We also calculated the prevalence of CMD, defined by reduced Coronary Flow Reserve (CFR <2.5), and calculated the prevalence of low BEW and low DMVC (values below the 25^th^ percentile) in each group.

**Results:** The three study groups were defined as having 37-53; 54-66 and 67-77 years of age, respectively. Oldest (3^rd^ tertile) patients show lower hyperemic flow velocity (46.7±14.4 vs 45.1±12.4 vs 38.4±11.5 cm.s-1, p=0.019), lower DMVC (1.90±0.71 vs 1.44±0.56 vs 1.37±0.67 cm.s^-1^.mmHg^-1^, p<0.001) and lower BEW intensity (5.9 [2.9-8.4] vs 4.8 [2.9-6.8] vs 4.4 [3.4-6.3] x10^6^ W.m^-2^.s^-1^, p=0.094). Older age was also found to be an independent predictor of lower cumulative BEW intensity (B −0.10, 95% CI: −0.17 to −0.09, p=0.021) and lower DMVC (B −0.25 95% CI: −0.45 to −0.09, p=0.027). In patients with CMD as determined by CFR <2.5, the prevalence of BEW intensity and DMVC below the 25^th^ percentile increased with age (25.0%, vs 52.0% vs 72.7%, for the 1^st^, 2^nd^ and 3^rd^ age tertiles, respectively, p=0.010).

**Conclusions:** Ageing is independently associated with structural microcirculatory remodelling that is reflected in BEW intensity and DMVC measurements and increased prevalence of structural CMD. These results are important in understanding non-obstructive mechanisms of myocardial ischemia in the elderly.

**CLINICAL PERSPECTIVE:** **What is new?**

- Ageing is independently associated with structural remodelling of the coronary microcirculation.
- Coronary structural microcirculatory remodelling is reflected by reductions in the Backward Expansion Wave intensity and in Diastolic Microvascular Conductance.

**What are the clinical implications?**

- lder patients exhibit lower hyperemic flow response to pharmacological hyperemia.
- can explain the observed age-related differences between hyperemic and non-hyperemic indices of functional stenosis classification.

## INTRODUCTION

Emerging evidence suggests the presence of age-related microcirculatory dysfunction. This phenomenon is characterized by dampened myocardial blood flow increase in response to adenosine and increased microvascular resistance on invasive coronary physiology studies[1,2]

Coronary microcirculatory dysfunction (CMD) is a complex pathophysiological phenomena that can obey to functional or structural derangements[3,4] Structural remodelling of the microcirculation, which can be caused among other causes by reduced capillary density (rarefaction), inward hypertrophic arteriolar remodelling and periarteriolar fibrosis, leads to decreased microcirculatory conductance, myocardial ischemia and confers a worse long-term prognosis [5–7]

Several intracoronary physiology indices have been validated against pathological evidence of structural microvascular remodelling. Diastolic microvascular conductance (DMVC) can be derived from combined intracoronary Doppler velocity and pressure measurements. Previous studies have shown that this index reflects better the haemodynamic consequences of arteriolar obliteration and capillary rarefaction than coronary flow reserve (CFR) or whole-cycle microcirculatory resistance[8]. The Backward Expansion Wave (BEW), an index derived from wave intensity analysis (WIA), constitutes a surrogate for increase in microvascular capacitance over early diastole, which is largely determined by capillary density[9–11]

As the fundamental pathological mechanisms of age-induced CMD remain largely unknown, in this study, we sought to comprehensively assess the pathophysiology underlying age-related microcirculatory dysfunction using WIA, DMVC and invasive indices of coronary physiology.

## METHODS

### Study Population

This study is an analysis of the Iberian-Dutch-English (IDEAL) registry, which is an international, multicenter registry that contains prospective collected data from combined pressure and Doppler flow velocity measurements from patients with stable angina who were treated at three participating centers: Amsterdam Medical Center in the Netherlands, Imperial College in the United Kingdom and Hospital Clínico San Carlos in Spain[12]. All enrolled patients were scheduled to undergo elective coronary angiography with physiological interrogation and informed consent for acquisition of additional physiological data for research purposes was obtained in all patients. The methodology for acquiring physiological data was standardized across all participating centers. Our study focused on patients without epicardial coronary stenosis on angiography and without diabetes mellitus, the latter a condition associated with both structural and functional derangements of the microcirculation that might impede a clear outlining of the relationship between age and structural microvascular remodelling [13,14]. Additional details regarding the inclusion and exclusion criteria can be found in the supplementary appendix of the original study[12].

### Coronary Angiography Intracoronary physiology

Coronary angiography and pressure-flow assessments were conducted according to standard protocols. Following the administration of intracoronary nitrates (100mcg or 200mcg), angiographic views were obtained and quantitative coronary angiography (QCA) was performed using validated software (CAAS II, Pie Medical Imaging, Maastricht, the Netherlands). Physiology measurements were taken after coronary angiography. A ComboWire XT guidewire(Philips Volcano, San Diego, USA) that combines pressure and Doppler flow velocity measurements was advanced into the distal target coronary artery after pressure equalization at the coronary ostium. Patients underwent wire-based recordings of mean aortic pressure (Pa), mean distal intracoronary pressure (Pd) and Doppler-flow velocities (resting and hyperemic flow by average peak velocity in mL/min). Stable hyperemia was achieved using intravenous adenosine perfusion at 140mcg/Kg/min for a minimum of 2 minutes, or by intracoronary bolus injection. A drift check was performed and considered acceptable if it was ≤0.02. All electrocardiogram, pressure and flow velocity data were extracted from the device console (ComboMap, Volcano Corporation, San Diego, CA, USA).

Several physiological indices were derived from the collected data. CFR was calculated as the ratio of the hyperemic averaged peak flow velocity (APV) to that during rest in mL/min[15]. HMR was calculated as the ratio between hyperemic Pd to hyperemic APV and expressed as mmHg.cm^-1^.s^-1^[16]. DMVC, which has also been described as Instantaneous Hyperaemic Diastolic Velocity Pressure Slope, was defined as the slope (beta coefficient) of the relationship between hyperemic Pd and AVP flow in mid to end diastole and was represented by a regression line (y= a + bx), expressed in cm.s^-1^.mmHg^-1^[17].

Wave Intensity is a measure of energy per area carried by arterial waves that travel through the cardiovascular system and is obtained by analysing the phasic changes in local pressure and flow velocity[9]. A dedicated software described elsewhere was used for calculations[11]. The recordings of pressure and Doppler flow velocity were filtered using the Savitzky-Golay method[11,18] and analysed for 30 cardiac cycles. The changes in pressure were separated into wave components that originated from the proximal vessel or the coronary microcirculation. The speed of the wave was estimated using the sum of squares method, assuming a blood density of 1050Kg.m^-3^ [19].

We assessed a total of six intracoronary waves, including three proximal (aortic or forward) waves and three distal (myocardial or backward) originating waves (Figure 1). The first forward wave occurring during ventricular systole, after the opening of the aortic valve, was defined as the Forward Compression Wave (FCW). The slowing of ventricular contraction during end-systole created the Forward Expansion Wave (FEW), which had a suction effect applied from the proximal coronary tree. The FCW has an identifiable second peak that originates when the aortic valve closes and creates a small, but measurable compressing wave with an accelerating effect on coronary blood flow. The early Backward Compression Wave (BCW) is created by ventricular isovolumetric contraction, while a late BCW is created during early systole as the sum of sustained compression of the microcirculation and as a reflection of the FCW from distal reflecting sites in the coronary three. Finally, the BEW, the most clinically relevant wave, originates at the end of the systole and during isovolumetric relaxation and is created by the re-expansion of the compressed intra-myocardial arteries with a sudden drop in resistance in the coronary microcirculation. We calculated the energy (W.m^-2^/s^2^.10^5^) from cumulative (area under the curve) BEW (C-BEW) and compared them between groups. To simplify interpretation, BEW values were presented as positive numbers. We also calculated the BEW energy fraction (BEW-EF), defined as the cumulative wave intensity divided by its integral over the cardiac period, which represents the fraction of the BEW energy relative to the net wave energy generated throughout the cardiac cycle.

**FIGURE 1.** Wave intensity profile in an obstructed coronary artery. (Upper panel): waves originating from both the proximal (positive values - red) and distal circulation (negative values – green). Six waves are present but the BEW is the main determinant of coronary flow. (Lower panel): Wave intensity can explain the complex pressure-flow relationship in the coronary artery throughout the cardiac cycle, showing reductions in pressure in increments in coronary flow during diastole due to the decompression of the microcirculation (From Sen S, Petraco R, Mayet J and Davies J. Wave intensity analysis in the human coronary circulation in health and disease. Current Cardiol Rev. 2014; 10:17-23; with permission).

### Age and Physiology Indices Relationship

To assess the impact of age on intracoronary physiology indices, we stratified our patient cohort into three groups based on age tertiles. We then evaluated correlations between these indices and age. Additionally, we calculated the C-BEW and compared it between the different age groups. Finally, we calculated the prevalence of CMD, defined by reduced Coronary Flow Reserve (CFR <2.5) in the absence of epicardial lesions, and calculated the prevalence of low BEW and low DMVC (as determined by values below the 25^th^ percentile) in each group. As a supplementary analysis, we also studied the relationship between BEW and structural CMD endotype as defined according to Rahman et al. [3].

### Statistical Analysis

Continuous variables that followed a normal distribution were reported as mean and standard deviation, while categorical variables were presented as absolute counts an percentages. We calculated 95% confidence intervals (CIs) for means of continuous variables and percentages of categorical variables were calculated using t-tests and Clopper-Pearson (Exact) approaches, respectively. To adjust for potential confounders, including interrogated target vessel with LAD artery lesion, presence of coronary artery disease risk factors and number of interrogated vessels per patient, we calculated adjusted correlations between coronary physiology indices and age. For variables that followed a normal distribution, we used t-tests, one-way ANOVA test and Tukey’s post-hoc analysis were used to compare means. Variable with non-normal distribution were compared using Mann-Whitney U tests. The Chi-square test was used to compare prevalences between different groups. We applied linear regression analysis to predict BEW intensity based on a broad range of admission parameters. To correct for possible unknown effects between more than one vessel interrogated per patient, we used generalized estimation equations (GEEs). All statistical analyses were performed using commercially available software (SPSS 28.0, IBM). Statistical significance was defined as a bilateral p-value <0.05.

## RESULTS

### Study Population

Table 1 shows the clinical and angiographic characteristics of the study population. We stratified the total population (99 patients and 165 vessels) into tertiles based on age resulting in the following groups: 1^st^ tertile: 37-53 years; 2^nd^ tertile: 54-66 years; and 3^rd^ tertile: 67-77 years. We observed a higher proportion of male gender and active smoking status in the younger age groups (p=0.029 and p=0.008, respectively), while the prevalence of hypertension was higher in the older age groups (p=0.010).

**Table 1:** General Characteristics of the Study Population.

|  | Overall<br>(n=99) | [37-53] years<br>(n=33) | [54-66] years<br>(n=29) | [67-77] years<br>(n=37) | p-value |
| --- | --- | --- | --- | --- | --- |
| **Baseline Demographics** |  |  |  |  |  |
| Age (years) | 57.7 ± 8.9 | 48.2 ± 3.8 | 56.7 ± 2.3 | 66.9 ± 5.0 | - |
| Male | 60.6 | 78.8 | 48.3 | 54.1 | 0.029 |
| BMI (Kg/m<sup>2</sup>) | 26.9 ± 3.6 | 26.3 ± 3.8 | 28.7 ± 3.9 | 26.3 ± 3.0 | 0.064 |
| **Medical History** |  |  |  |  |  |
| Prior MI | 4.0 | 9.1 | 3.4 | 0.0 | 0.153 |
| Hypertension | 43.4 | 27.3 | 37.9 | 62.2 | 0.010 |
| Dyslipidaemia | 51.5 | 51.5 | 62.1 | 43.2 | 0.316 |
| Smoker | 36.4 | 57.6 | 27.6 | 24.3 | 0.008 |
| **Current Medication** |  |  |  |  |  |
| Betablocker | 50.5 | 51.5 | 51.7 | 48.6 | 0.960 |
| Statin | 59.6 | 63.6 | 65.5 | 51.4 | 0.429 |
| ACE inhibitor | 12.1 | 6.1 | 6.9 | 12.1 | 0.081 |
| Calcium Channel Blocker | 16.2 | 12.1 | 24.1 | 13.5 | 0.377 |
| Acetyl Salicylic Acid | 69.7 | 69.7 | 65.5 | 73.0 | 0.807 |

BMI: body mass index; MI: myocardial infarction; ACE: angiotensin converting enzyme

### Haemodynamic parameters

Table 2 shows that in the interrogated vessel population (n=165) there were no significant differences between the groups in terms of the interrogated coronary artery or the number of interrogated arteries per patient. We found a positive linear correlation between C-BEW and BEW-EF (r^2^=0.35, 95% CI: 0.11 to 0.48, p=0.012). C-BEW also correlated positively with DMVC (r^2^=0.21, 95% CI: 0.07 to 0.37, p=0.040) and negatively with HMR (r^2^=-0.23, 95% CI: −0.40 to −0.10, p=0.023). These findings reinforce the intrinsic direct relationship between capillary density, microcirculatory conductance and resistance.

**Table 2.** Invasive physiological assessment.

|  | Population<br>(n=99,<br>vessels=165) | [37-53] years<br>(n=33,<br>vessels=51) | [54-66] years<br>(n=29,<br>vessels=66) | [67-77] years<br>(n=37,<br>vessels=48) | p-value |
| --- | --- | --- | --- | --- | --- |
| **Interrogated Artery** |  |  |  |  |  |
| Right coronary | 24.8 | 19.6 | 24.2 | 25.0 | 0.282 |
| Left anterior descending | 40.0 | 47.1 | 39.4 | 39.6 | 0.574 |
| Left circumflex | 35.2 | 33.3 | 36.4 | 35.4 | 0.947 |
| Vessels per patient | 1 [1-2] | 1 [1-2] | 2 [1-2] | 1 [1-2] | 0.352 |
| **Physiological and Angiographic indices** |  |  |  |  |  |
| Basal Pd/Pa | 0.98 ± 0.03 | 0.98 ± 0.02 | 0.98 ± 0.02 | 0.99 ± 0.03 | 0.069 |
| Baseline flow velocity (cm.s<sup>-1</sup>) | 17.2 ± 6.15 | 18.5 ± 6.6 | 17.0 ± 6.3 | 16.6 ± 4.4 | 0.118 |
| Hyperemic flow velocity (cm.s<sup>-1</sup>) | 44.6 ± 15.7 | 46.7 ± 14.4 | 45.1 ± 12.4 | 38.4 ± 11.5 | 0.019 |
| CFR | 2.68 ± 0.79 | 2.67 ± 0.80 | 2.70 ± 0.81 | 2.58 ± 0.73 | 0.062 |
| HMR | 2.21 ± 0.80 | 1.92 ± 0.49 | 2.60 ± 0.74 | 2.70 ± 0.81 | 0.002 |
| BMR | 6.24 ± 2.33 | 5.62 ± 1.83 | 6.41 ± 2.65 | 6.89 ± 1.88 | 0.040 |
| DMVC | 1.57 ± 0.66 | 1.90 ± 0.71 | 1.44 ± 0.56 | 1.37 ± 0.67 | <0.001 |
| Baseline flow velocity (cm.s<sup>-1</sup>) | 17.2 ± 6.2 | 18.5 ± 6.6 | 17.0 ± 6.3 | 16.6 ± 4.4 | 0.118 |
| Hyperemic flow velocity (cm.s<sup>-1</sup>) | 44.6 ± 15.7 | 46.7 ± 14.4 | 45.1 ± 12.4 | 38.4 ± 11.5 | 0.019 |
| **Wave Intensity Analysis** |  |  |  |  |  |
| Cumulative BEW (x10<sup>6</sup> W.m<sup>-2</sup>.s<sup>-1</sup>) | 4.9 [3.4-7.6] | 5.9 [2.9-8.4] | 4.8 [2.9-6.8] | 4.4 [3.4-6.3] | 0.094 |

Pd: distal pressure; Pa: aortic pressure; FFR: fractional flow reserve; iFR: instantaneous wave-free ratio; CFR: coronary flow reserve; HMR: hyperemic microvascular resistance; basal microvascular resistance; IHDVPS: instantaneous hyperemic diastolic velocity pressure slope; BEW: backward expansion wave

### Effect of age on pressure and flow based functional indices

A significant reduction of hyperaemic flow velocity was noted in older patients (p=0.019 – Table 2). Age correlated negatively with C-BEW intensity (r^2^=-0.13, 95% CI −0.27 to −0.05, p=0.037 – Figure 2A), DMVC (r^2^=-0.35, 95% CI: −0.48 to −0.17, p<0.001 – Figure 2B) and CFR (r^2^=-0.16, 95% CI −0.35 to −0.07, p=0.033 – Figure 2C) and positively with HMR (r^2^=0.31, 95% CI: 0.19 to 0.41, p<0.001 −Figure 2D). Furthermore, older patients had a significant higher prevalence of increased microvascular dysfunction as defined by HMR≥2.5 (11.8% vs 24.4% vs 39.3% for 1^st^, 2^nd^ and 3^rd^ age tertiles respectively, p= 0.019).

**FIGURE 2.** Age correlation with microcirculatory physiology indices. A – Age correlation with C-BEW: B – Age correlation with DMVC; C – Age correlation with CFR; D – Age correlation with HMR. C-BEW – Cumulative Backward Expansion Wave; DMVC – Diastolic Microvascular Conductance, CFR – Coronary Flow Reserve; HMR – Hyperemic Microvascular Resistance.

### Effect of age on Backward Expansion Wave Intensity

C-BEW intensity was compared between the age tertiles, and lower intensity values were observed in the older age groups, although statistical significance was not reached (p=0.094). The same trend was observed regarding BEW-EF (p=0.085) - Table 3. However, after adjusting for demographic (gender, hypertension, smoking status, body mass index, dyslipidaemia) and angiography (target vessel and number of interrogated vessels per patient) related factors, we found that age was a significant and independent predictor of lower cumulative BEW intensity (p=0.030) - Table 4.

**Table 3.** Wave Intensity Analysis for each wave component.

| | Cumulative BEW ($\times 10^5 \text{ W.m}^{-2}.\text{s}^{-1}$) | BEW energy fraction (%) |
| --- | --- | --- |

|  | Population<br>(n=99, vessels=165) | [37-53] years<br>(n=33, vessels=51) | [54-66] years<br>(n=29, vessels=66) | [67-77] years<br>(n=37, vessels=48) | p-value |
| --- | --- | --- | --- | --- | --- |
| **Wave intensity analysis** |  |  |  |  |  |
| Cumulative FCW ($\times 10^6 \text{ W.m}^{-2}.\text{s}^{-1}$) | 4.5 [3.1-10.1] | 6.7 [3.6-10.5] | 4.7 [3.0-7.6] | 3.9 [2.3-8.3] | 0.995 |
| Cumulative FCW 2<sup>nd</sup> peak ($\times 10^6 \text{ W.m}^{-2}.\text{s}^{-1}$) | 2.3 [1.0-4.9] | 1.3 [0.7-4.5] | 1.3 [0.8-3.1] | 0.7 [0.3-1.8] | 0.648 |
| Cumulative FEW ($\times 10^6 \text{ W.m}^{-2}.\text{s}^{-1}$) | 2.4 [0.8-5.7] | 1.5 [0.5-6.5] | 2.1 [0.7-5.9] | 2.8 [1.9-5.8] | 0.778 |
| Cumulative early BCW ($\times 10^6 \text{ W.m}^{-2}.\text{s}^{-1}$) | 0.5 [0.2-2.1] | 1.1 [0.2-2.4] | 1.6 [0.5-2.7] | 0.9 [0.5-3.4] | 0.622 |
| Cumulative late BCW ($\times 10^6 \text{ W.m}^{-2}.\text{s}^{-1}$) | 2.4 [1.4-5.6] | 1.7 [0.9-2.9] | 2.3 [1.4-4.6] | 2.0 [0.7-4.4] | 0.534 |
| Cumulative BEW ($\times 10^6 \text{ W.m}^{-2}.\text{s}^{-1}$) | 4.9 [3.4-7.6] | 5.9 [2.9-8.4] | 4.8 [2.9-6.8] | 4.4 [3.4-6.3] | 0.094 |
| BEW energy fraction (%) | 32.7 ± 6.8 | 35.9 ± 9.7 | 31.3 ± 11.2 | 29.2 ± 9.9 | 0.085 |

FCW: forward compression wave; FEW: forward compression wave; BCW: backward compression wave; BEW: backward expansion wave.

**Table 4.** Estimating Model for BEW energy fraction Generalized Equations Linear intensity and prediction.

|  | B (x10<sup>6</sup>) | 95% CI (x10<sup>6</sup>) | p-value | B | 95% CI | p-value |
| --- | --- | --- | --- | --- | --- | --- |
| Intercept | 107.1 | - | - | 19.2 | - | - |
| Age (years) | -0.41 | -0.71 to -0.20 | 0.030 | -1.1 | -2.8 to 0.1 | 0.117 |
| Male gender | 0.95 | -19.5 to 21.4 | 0.927 | 2.1 | -3.7 to 5.4 | 0.915 |
| Hypertension | 12.9 | 9.8 to 16.2 | <0.001 | 4.5 | -0.7 to 7.4 | 0.216 |
| Dyslipidaemia | 8.2 | -5.5 to 7.1 | 0.800 | 1.7 | -2.2 to 3.9 | 0.405 |
| Smoker | 8.9 | -13.5 to 31.4 | 0.433 | 3.6 | -1.2 to 6.9 | 0.377 |
| BMI | -0.12 | -0.81 to 0.6 | 0.727 | 0.2 | -0.5 to 1.6 | 0.518 |
| Betablocker | 8.8 | 3.4 to 14.3 | 0.002 | 7.4 | 4.2 to 16.0 | 0.003 |
| ACE inhibitor | 20.6 | -10.8 to 51.3 | 0.187 | 5.3 | -5.5 to 12.9 | 0.399 |
| Calcium Chanel Blocker | -6.7 | -23.7 to 10.4 | 0.443 | 9.7 | -10.1 to 21.7 | 0.894 |
| Statin | 6.9 | -10.1 to 24.6 | 0.445 | 1.2 | -7.1 to 5.6 | 0.603 |
| Left Coronary Artery | 11.9 | 5.1 to 18.7 | <0.001 | 6.7 | 1.9 to 11.3 | <0.001 |

### Age and CMD mechanisms

Figure 3A illustrates the underlying mechanisms of age-related CMD. In this population, the prevalence of CMD, as determined by CFR < 2.5, was found in 52.5% of patients (n=52). We also found a higher proportion of patients with reduced CFR in older patients, although without statistical significance (48.5% vs 51.0% vs 64.7%, for the 1^st^, 2^nd^ and 3^rd^ age tertiles respectively, p=0.129). However, the proportion of patients with CMD whose C-BEW or DMVC values are below the 25^th^ percentile increased significantly with age (25.0%, vs 52.0% vs 72.7%, for the 1^st^, 2^nd^ and 3^rd^ age tertiles, respectively, p=0.010). This suggests that CMD in the elderly is determined predominantly by structural derangement of the coronary microcirculation in opposition to younger patients. In contrast, in the absence of CMD (47.5%, n=47), no differences were found in the proportion of patient with low C-BEW or DMVC values between age groups (23.5% vs 29.4% vs 33.3%, for the 1^st^, 2^nd^ and 3^rd^ age tertiles respectively, p=0.961 – Figure 3B).

**FIGURE 3.** Relationship between low BEW and low DMVC in age-related CMD. A - The prevalence of CMD as determined by CFR < 2.5 increases with age. However, unlike younger patients, CMD in the elderly is associated with higher prevalence of structural remodeling, as showed by higher proportion of patients with BEW and DMVC values below the 25^th^ percentile. B – In patients with normal CFR, the proportion of patients with BEW and DMVC values below the 25^th^ percentile does not vary significantly. C-BEW – Cumulative Backward Expansion Wave, DMVC – Diastolic Microvascular Conductance, CMD – Coronary Microcirculatory Dysfunction

## DISCUSSION

In our study we hypothesized that structural remodelling is the main pathophysiological mechanism for the development of age-related CMD. To test our hypothesis, we used WIA and DMVC as diagnostic tools in a cohort of non-diabetic patients with chronic coronary syndromes without angiographic epicardial lesions. We examined age-related changes in C-BEW intensity and DMVC, their influence on invasive indices of coronary physiology and in the prevalence of structural CMD.

While there is growing awareness that myocardial ischaemia in chronic coronary syndromes may result from obstructive and non-obstructive causes[20,21], there is limited evidence on the effect of age on microcirculatory function, a knowledge gap that requires urgent attention due to the growing number of elderly people worldwide. Our findings support that older age is independently associated with significant reductions in C-BEW intensity and DMVC, supporting the existence of age-induced structural remodelling of the coronary microcirculation (Central Illustration).

**CENTRAL ILLUSTRATION.** Age-related microvascular remodeling and its impact on microvascular expansion and conductance Upper Panel – From left to right – Schematic representation of progressive microvascular structural remodeling and rarefaction associated with age; Middle Panel – Boxplot graphic for cumulative BEW for each age strata (p-value obtained from linear regression GEE); Lower Panel – Boxplot graphic for MVC for each age strata C-BEW – Cumulative Backward Expansion Wave, DMVC – Diastolic Microvascular Conductance, GEE – Generalized Estimation Equation.

The pathophysiology of CMD is complex and multifactorial, involving both functional and structural mechanisms[4]. The BEW (also known as backward decompression or “suction” wave), caused by the rapid re-expansion of the capillary bed during early-diastole after being compressed during systole, is the largest contributor to coronary blood flow[9]. Structural changes of the microcirculation, such as perivascular fibrosis, arteriolar thickening and capillary rarefaction can decrease BEW intensity, as previously shown in patients with hypertrophic cardiomyopathy, aortic stenosis, myocardial infarction, heart failure and cardiac allograft patients[10,22–24]. Our study demonstrates that ageing is also independently associated with this structural remodelling phenomena.

Previous research by our group has demonstrated that ageing is associated with a decrease in the microcirculatory vasodilatory response to adenosine administration. This leads to an increase in functional stenosis classification discrepancies between hyperaemic and non- hyperaemic indices, as reflected by an age-dependent increase in Fractional Flow Reserve (FFR) values[1,25]. Other researchers have also described the impact of other age-related modifications in the coronary microcirculation, which can further explain our findings[26–28] Nevertheless, our current study provides invasive physiology data that adds knowledge to the understanding of the pathophysiology of age-induced reduction hyperaemic blood flow, particularly in patients older than 60 years [29].

Recently, a dichotomous classification of CMD as having a structural or functional cause has been proposed by Rahman et al. based on combined measurements of Doppler-based CFR and whole-cycle microcirculatory resistance (HMR)[3]. These authors propose, based on CFR and HMR values, the existence of either functional CMD (CFR<2.5 and HMR <2.5 mmHg.cm^-1^.s^-1^) and structural (CFR<2.5 and HMR ≥2.5 mmHg.cm^-1^.s^-1^) CMD endotypes. We applied the same dichotomous classification to our population and found that patients with structural CMD were older (64.6 ± 7.1 vs 56.7 ± 8.4 years, p<0.001) and that its increased progressively across age tertiles (3.0% vs 16.3% vs 41.2% for the 1^st^, 2^nd^ and 3^rd^ age tertiles respectively, p=0.002). At a difference, patients with functional CMD were younger (53.4 ± 8.7 years vs 59.4 ± 8.2 years, p=0.049) and its prevalence decreased across age tertiles (45.5% vs 34.7% vs 23.5% for the 1^st^, 2^nd^ and 3^rd^ age tertiles respectively, p=0.029) – Supplementary Figure 1. In addition, we also constructed a GEE binary logistic model to identify predictors of structural CMD and found an independent association with older age (p<0.001), lower C-BEW intensity (p<0.001), lower BEW-EF (p=0.009) and lower DMVC (p=0.041) – Supplementary Table 1.

Hence, our study reveals that the prevalence of structural CMD increases with age, while the prevalence of functional CMD is higher in younger patients. This finding adds evidence to support the concept that functional CMD could represent an early stage of the disease, as characterized by endothelial dysfunction, microvascular spasm, and transient microcirculatory impairment. With sustained chronic inflammation, microvascular injury and maladaptation, it can evolve to a structural CMD endotype, hallmarked by perivascular fibrosis, capillary rarefaction and irreversible tissue damage, and raising awareness for the importance of early detection and management to prevent disease progression[30]. Finally, our study also suggests that age-related reductions in BEW intensity and DMVC could be used as surrogates to predict the development of structural CMD in ageing populations.

## STRNGTHS AND LIMITATIONS OF THE STUDY

We believe that our study has several methodological strengths, including 1) the use of combined intracoronary Doppler and pressure, a powerful combo that allows analysis of specific segments of the cardiac cycle such as diastolic microvascular conductance and wave intensity analysis; 2) using as a standard of reference physiology indices that have been validated against histological changes in microcirculatory structure in endomyocardial biopsies; 3) restricting the analyses to patients without epicardial stenosis which might modulate the status of the subtended microcirculation[31]; and 4) excluding patients with diabetes mellitus due to its effect on microvascular structure [32]. As limitations we must highlight that this is a subgroup analysis of the IDEAL registry[12], and that therefore we cannot rule out the effect of selection bias and confounding variables. Of note, patients in the IDEAL registry did not undergo coronary acetylcholine testing, and therefore the status of endothelium-dependent pathway remains unknown.

## CONCLUSIONS

Ageing is associated with structural remodelling of the coronary microcirculation manifested by reductions in BEW intensity and DMVC that are associated with higher prevalence of structural CMD. These findings contribute to explain the lower hyperemic flow found in older populations and the observed age-related differences between hyperaemic and non- hyperaemic indices of functional stenosis classification.

Conflicts of Interest:

Dr. Lombardi has served as speaker for Phillips

Dr. Travieso has received unconditional educational grants from Philips.

Dr. Nijjer has served as speaker and / or advisory board member for Philips. Dr. Piek has served as speaker and / or advisory board member for Philips. Dr. van de Hoef is a consultant for Philips

Dr. Echarravía-Pinto has received speaker fees from Boston Scientific

Dr. Mejía-Renteria served as speaker at educational events organised by Abbott, Boston Scientific and Philips Healthcare

Dr. Davies holds IP pertaining to iFR technology and is also a consultant and recipient of research funding from Philips Volcano.

Dr. Escaned has served as speaker and / or advisory board member for Abbott, Boston Scientific and Philips.

Dr. Faria, Dr. van der Hoeven, Dr. Heemelaar, Dr. de Waard, Dr. Sen, Dr, van de Hoef, Dr. Petraco and Dr. Niels van Royen have nothing to declare.

## Data Availability

The data that support the findings of this study are available on request from the corresponding author (JE)

## LIST OF ABREVIATIONS

APV: Average Peak Flow Velocity
BCW: Backward Compression Wave
BEW: Backward Expansion Wave
BEW- EF: Backward Expansion Wave Energy Fraction
C-BEW: Cumulative Backward Expansion Wave
CFR: Coronary Flow Reserve
CMD: Coronary Microcirculatory Dysfunction
DMVC: Diastolic Microvascular Conductance
IDEAL: Iberian-Dutch-English
FCW: Forward Compression Wave
FEW: Forward Expression Wave
FFR: Fractional Flow Reserve
HMR: Hyperemic Microvascular Conductance
Pa: mean aortic pressure
Pd: mean distal coronary pressure
QCA: Quantitative Coronary Angiography
WIA: Wave Intensity Analysis

## REFERENCES

1. Faria DC, Lee JM, Van Der Hoef T, Mejía-Rentería H, Echavarría-Pinto M, Baptista SB, Cerrato E, García-García H, Davies J, Onuma Y, et al. Age and functional relevance of coronary stenosis: a post hoc analysis of the ADVISE II trial. EuroIntervention. 2021;17:757–64.

2. Mejia-Renteria H, Faria D, Lee JM, Lee SH, Jung JH, Doh JH, Nam CW, Shin ES, Hoshino M, Sugiyama T, et al. Association between patient age, microcirculation, and coronary stenosis assessment with fractional flow reserve and instantaneous wave-free ratio. Catheter Cardiovasc Interv. 2022;99.

3. Rahman H, Ryan M, Lumley M, Modi B, Mcconkey H, Ellis H, Scannell C, Clapp B, Marber M, et al. Coronary Microvascular Dysfunction Is Associated With Myocardial Ischemia and Abnormal Coronary Perfusion During Exercise. Circulation. 2019;140:1805.

4. Mejía-Rentería H, van der Hoeven N, van de Hoef TP, Heemelaar J, Ryan N, Lerman A, van Royen N, Escaned J. Targeting the dominant mechanism of coronary microvascular dysfunction with intracoronary physiology tests. Int J Cardiovasc Imaging. 2017;33:1041–59.

5. Mohammed SF, Hussain S, Mirzoyev SA, Edwards WD, Maleszewski JJ, Redfield MM. Coronary Microvascular Rarefaction and Myocardial Fibrosis in Heart Failure with Preserved Ejection Fraction. Circulation. 2015;131:550.

6. Bajaj NS, Singh A, Zhou W, Gupta A, Fujikura K, Byrne C, Harms HJ, Osborne MT, Bravo P, Andrikopolou E, et al. Coronary Microvascular Dysfunction, Left Ventricular Remodeling, and Clinical Outcomes in Patients with Chronic Kidney Impairment. Circulation. 2020:21–33.

7. Broyd CJ, Echavarria-Pinto M, Cerrato E, Escaned J. Evaluation of Microvascular Disease and Clinical Outcomes. Interv Cardiol Clin. 2015;4:443–57.

8. Escaned J, Flores A, García-Pavía P, Segovia J, Jimenez J, Aragoncillo P, Salas C, Alfonso F, Hernández R, Angiolillo DJ, et al. Assessment of microcirculatory remodeling with intracoronary flow velocity and pressure measurements: validation with endomyocardial sampling in cardiac allografts. Circulation. 2009;120:1561–8.

9. Broyd CJ, Davies JE, Escaned JE, Hughes A, Parker K. Wave intensity analysis and its application to the coronary circulation. Glob Cardiol Sci Pract. 2017;2017.

10. Broyd CJ, Hernández-Pérez F, Segovia J, Echavarría-Pinto M, Quirós-Carretero A, Salas C, Gonzalo N, Jiménez-Quevedo P, Nombela-Franco L, Salinas P, et al.. Identification of capillary rarefaction using intracoronary wave intensity analysis with resultant prognostic implications for cardiac allograft patients. Eur Heart J. 2018;39:1807–14.

11. Davies JE, Whinnett ZI, Francis DP, Manisty CH, Aguado-Sierra J, Willson K, Foale RA, Malik IS, Hughes AD, Parker KH, et al. Evidence of a dominant backward-propagating ‘suction’ wave responsible for diastolic coronary filling in humans, attenuated in left ventricular hypertrophy. Circulation. 2006;113:1768–78.

12. Nijjer SS, de Waard GA, Sen S, van de Hoef TP, Petraco R, Echavarría-Pinto M, van Lavieren MA, Meuwissen M, Danad I, Knaapen P, et al. Coronary pressure and flow relationships in humans: phasic analysis of normal and pathological vessels and the implications for stenosis assessment: a report from the Iberian-Dutch-English (IDEAL) collaborators. Eur Heart J. 2016;37:2069–80.

13. Sezer M, Kocaaga M, Aslanger E, Atici A, Demirkiran A, Bugra Z, Umman S, Umman B. Bimodal Pattern of Coronary Microvascular Involvement in Diabetes Mellitus. J Am Hear Assoc Cardiovasc Cerebrovasc Dis. 2016;5.

14. Leung M, Biostat M, Leung DY. Coronary microvascular function in patients with type 2 diabetes mellitus. EuroIntervention. 2016;11:1111–7.

15. Fearon WF, Farouque HMO, Balsam LB, Cooke DT, Robbins RC, Fitzgerald PJ, Yeung AC, Yock PG. Comparison of Coronary Thermodilution and Doppler Velocity for Assessing Coronary Flow Reserve. Circulation. 2003;108:2198–200.

16. de Waard GA, Nijjer SS, van Lavieren MA, van der Hoeven NW, Petraco R, van de Hoef TP, Echavarria-Pinto M, Sen S, van de Ven PM, Knaapen P, et al. Invasive minimal Microvascular Resistance is a New Index to Assess Microcirculatory Function Independent of Obstructive Coronary Artery Disease. J Am Heart Assoc. 2016;5.

17. van der Hoeven NW, de Waard GA, Quirós A, de Hoyos A, Broyd CJ, Nijjer SS, van de Hoef TP, Petraco R, Driessen RS, Mejía-Rentería H, et al. Comprehensive physiological evaluation of epicardial and microvascular coronary domains using vascular conductance and zero flow pressure. EuroIntervention. 2019;14:E1593–600.

18. Ladwiniec A, White PA, Nijjer SS, O’Sullivan M, West NEJ, Davies JE, Hoole SP. Diastolic Backward-Traveling Decompression (Suction) Wave Correlates with Simultaneously Acquired Indices of Diastolic Function and Is Reduced in Left Ventricular Stunning. Circ Cardiovasc Interv. 2016;9.

19. Parker KH. An introduction to wave intensity analysis. Med Biol Eng Comput. 2009;47:175–88.

20. Koller A, Laughlin MH, Cenko E, De Wit C, Tóth K, Bugiardini R, Trifunovits D, Vavlukis M, Manfrini O, Lelbach A, et al. Functional and structural adaptations of the coronary macro- and microvasculature to regular aerobic exercise by activation of physiological, cellular, and molecular mechanisms: ESC Working Group on Coronary Pathophysiology and Microcirculation position paper. Cardiovasc Res. 2022;118:357–71.

21. Kunadian V, Chieffo A, Camici PG, Berry C, Escaned J, Maas AHEM, Prescott E, Karam N, Appelman Y, Fraccaro C, et al. An EAPCI Expert Consensus Document on Ischaemia with Non-Obstructive Coronary Arteries in Collaboration with European Society of Cardiology Working Group on Coronary Pathophysiology & Microcirculation Endorsed by Coronary Vasomotor Disorders International Study Group. Eur Heart J. 2020;41:3504–20.

22. Rolandi MC, Wiegerinck EMA, Casadonte L, Yong ZY, Koch KT, Vis M, Piek JJ, Baan J, Spaan JAE, Siebes M. Transcatheter Replacement of Stenotic Aortic Valve Normalizes Cardiac-Coronary Interaction by Restoration of Systolic Coronary Flow Dynamics as Assessed by Wave Intensity Analysis. Circ Cardiovasc Interv. 2016;9.

23. Camici P, Chiriatti G, Lorenzoni R, Bellina RC, Gistri R, Italiani G, Parodi O, Salvadori PA, Nista N, Papi L, et al. Coronary vasodilation is impaired in both hypertrophied and nonhypertrophied myocardium of patients with hypertrophic cardiomyopathy: A study with nitrogen-13 ammonia and positron emission tomography. J Am Coll Cardiol. 1991;17:879–86.

24. Rolandi MC, Nolte F, van de Hoef TP, Remmelink M, Baan J, Piek JJ, Spaan JAE, Siebes M. Coronary wave intensity during the Valsalva manoeuvre in humans reflects altered intramural vessel compression responsible for extravascular resistance. J Physiol. 2012;590:4623.

25. Faria D, Mejia-Renteria H, Lee JM, Lee SH, Travieso A, Jung JH, Doh JH, Nam CW, Shin ES, Hoshino M, Sugiyama T, et al.. Age-related changes in the coronary microcirculation influencing the diagnostic performance of invasive pressure-based indices and long-term patient prognosis. Catheter Cardiovasc Interv. 2022.

26. Jenner TL, Mellick AS, Harrison GJ, Griffiths LR, Rose’Meyer RB. Age-related changes in cardiac adenosine receptor expression. Mech Ageing Dev. 2004;125:211–7.

27. Echavarría-Pinto M, Gonzalo N, Ibañez B, Petraco R, Jimenez-Quevedo P, Sen S, Nijjer S, Tarkin J, Alfonso F, Núñez-Gil IJ, et al. Low coronary microcirculatory resistance associated with profound hypotension during intravenous adenosine infusion implications for the functional assessment of coronary stenoses. Circ Cardiovasc Interv. 2014;7:35–42.

28. Heusch G. Editorial: Adenosine and maximum coronary vasodilation in humans: Myth and misconceptions in the assessment of coronary reserve. Basic Res Cardiol. 2010;105:1–5.

29. Lee JM, Hwang D, Park J, Tong Y, Koo BK. Physiologic mechanism of discordance between instantaneous wave-free ratio and fractional flow reserve: Insight from 13N- ammonium positron emission tomography. Int J Cardiol. 2017;243:91–4.

30. Crea F, Camici PG, Merz CNB. Coronary microvascular dysfunction: an update. Eur Heart J. 2014;35:1101–11.

31. Gould KL. Pressure-flow characteristics of coronary stenoses in unsedated dogs at rest and during coronary vasodilation. Circ Res. 1978;43:242–53.

32. Gallinoro E, Paolisso P, Bertolone DT, Belmonte M, Esposito G, Munhoz D, Sakai K, Bermpeis K, Ohashi H, Storozhenko T, et al. Endotypes of coronary microvascular dysfunction in patients with and without Diabetes Mellitus. Eur Heart J. 2023;44.

